# Correlations Between EEG Spectral Power and Cerebral Perfusion in Adults Undergoing Extracorporeal Membrane Oxygenation

**DOI:** 10.1101/2022.06.07.22276122

**Authors:** Imad R. Khan, Irfaan A. Dar, Thomas W. Johnson, Emily Loose, Yama Y. Xu, Esmeralda Santiago, Kelly L. Donohue, Mark A. Marinescu, Igor Gosev, Giovanni Schifitto, Regine Choe, Olga Selioutski

**Affiliations:** Department of Neurology, University of Rochester Medical Center, Rochester, NY, USA; Department of Biomedical Engineering, University of Rochester, Rochester, NY, USA; School of Arts and Sciences, University of Rochester, Rochester, NY, USA; Department of Medicine, University of Rochester Medical Center, Rochester, NY, USA; Division of Cardiac Surgery, University of Rochester Medical Center, Rochester, NY, USA; Department of Electrical and Computer Engineering, University of Rochester, Rochester, NY, USA

**Keywords:** Hypoxic ischemic brain injury, coma, quantitative EEG, cerebral blood flow, neuromonitoring, neurovascular unit

## Abstract

**Objective:** Patients with acute lung or heart failure can experience hypoxic ischemic brain injury resulting in neurovascular unit (NVU) dysfunction. The NVU couples brain activity and perfusion. Extracorporeal membrane oxygenation (ECMO) is used for refractory lung and/or heart failure and often necessitates sedation, obfuscating neurological assessments. We hypothesized that combining cerebral electrographic activity and perfusion monitoring can detect brain injury in adults undergoing ECMO.

**Methods:** Fast Fourier transformation was used to identify fast (α+β) and slow (δ) power bands from cEEG. Diffuse correlation spectroscopy (DCS) measured blood flow index (BFI), a surrogate of perfusion, daily in adults undergoing ECMO. Correlations between α+β/δ ratio (ABDR) and BFI were compared between patients who were grouped into brain-injured and uninjured groups by neurologic exam and neuroimaging findings.

**Results:** Ten patients (21-78 years old, five females, five brain-injured) underwent cEEG and DCS monitoring. Sixty-eight monitoring sessions (average 127 minutes/session) were analyzed. ABDR-BFI correlation was significantly higher in uninjured patients than brain-injured ones. Sedation did not significantly impact CBF-power band correlation.

**Conclusions:** Brain-injured patients exhibited significantly less correlation between brain activity and perfusion, possibly as a result of NVU dysfunction.

**Significance:** ABDR-BFI correlation can be measured continuously and noninvasively at the bedside and may represent a marker of NVU dysfunction.

**Highlights:**

- Quantitative EEG and diffuse correlative spectroscopy can be used to measure markers of brain injury noninvasively, continuously, and at the bedside.
- Decreased ABDR-BFI correlation may be a marker of neurovascular decoupling in patients with hypoxic ischemic brain injury.
- ABDR-BFI correlation may be independent of analgosedation, which is used heavily in patients undergoing ECMO.

## 1. Introduction

Hypoxia and ischemia cause brain injury by damaging any of the components of the neurovascular unit (NVU), including neurons, glial cells, pericytes and/or endothelium. These cellular components communicate with one another via neurovascular coupling to ensure adequate blood flow to support neuronal function in response to metabolic demand. (Iadecola, 2017) Hypoxic-ischemic brain injury (HIBI) results in neuronal destruction, blood brain barrier disruption (Knowland et al., 2014), and formation of microvascular thrombi (Wada, 2017) amongst many other pathophysiological processes that may disrupt NVU function. Thus, the detection of NVU dysfunction holds potential as a biomarker of HIBI in critically-ill patients and provides basis for outcome prediction as well as targets for therapies aimed at preventing or mitigating neurologic injury after HIBI. On the other hand, demonstrating intact NVU function may assist in favorable outcome prognostication.

NVU function is commonly measured using blood oxygen level-dependent signals obtained by functional MRI, which highlights regional upregulations of perfusion in metabolically-active brain regions. (Raichle and Mintun, 2006) In addition to its assessment limited only to active brain regions, this modality offers low temporal resolution, making it unfeasible for continuous detection of NVU dysfunction in critically-ill populations. EEG assesses summated excitatory and inhibitory postsynaptic potentials of cortical neurons with excellent spatial and temporal resolution. (Kirschstein and Kohling, 2009) Recently, the combination of EEG and near-infrared spectroscopy (NIRS) has demonstrated the ability to detect NVD and predict MRI-confirmed brain injury in neonates with HIBI. (Das et al., 2021) By using near-infrared light, NIRS detects cerebral oxygen saturation, a surrogate for cerebral blood flow (CBF). Diffuse correlation spectroscopy (DCS) is a similar optical technology, however it directly measures CBF at the microvascular level. (Dar et al., 2020) DCS is a novel non-invasive technology that uses near-infrared light sources that penetrate through the skull, making it ideal for continuous monitoring of cortical blood flow at the bedside. DCS offers a quantitative blood flow index (BFI) proportional to microvascular blood flow. (Durduran and Yodh, 2014) Studies have demonstrated DCS can capture functional signals from brain cortex and successfully monitor cerebral blood flow during clinical interventions. (Buckley et al., 2013, Delgado-Mederos et al., 2018, Durduran and Yodh, 2014, Durduran et al., 2009, Kim et al., 2010, Mesquita et al., 2011)

Patients receiving extracorporeal membrane oxygenation (ECMO) therapy are critically ill with refractory cardiogenic shock (CS), cardiac arrest (CA), or acute respiratory distress syndrome (ARDS). This population is ideal to study a noninvasive, multimodal paradigm of NVD for a number of reasons. Up to 13% of ECMO patients with CS and CA and 1% of those with ARDS experience HIBI and presumably NVD, and mortality rates rise up to 89% in ECMO patients with brain injury. (Lorusso et al., 2016, Shoskes et al., 2020) ECMO patients often require high sedation to maintain ventilator compliance or prevent cannula dislodgement, obviating clinical neurologic examination. (Marhong et al., 2017) In lieu of the clinical exam, intracranial monitors can inform clinicians on cerebral physiology. However, placement is risky in ECMO patients and is not routinely performed due to coagulopathy from clotting factor deficiencies, platelet dysfunction, and anticoagulation to prevent circuit clotting. (Thomas et al., 2018) A noninvasive multimodal neuromonitoring paradigm is needed that combines perfusion and electrographic monitoring to inform the clinician of the existing degree of brain injury and identify patients at risk of secondary brain injury. This may ultimately provide a target for brain-focused resuscitation and prognostication. (Lorusso et al., 2017, Ong et al., 2021)

In this study, we evaluate the correlation of DCS with raw and quantitative EEG (qEEG) findings to infer NVU dysfunction in the cortex manifested as the absence of correlation between microvascular CBF and electrical neuronal activity. We hypothesized that BFI and EEG frequencies are consistently correlated in patients without brain injury and lacks correlation in adult ECMO patients with HIBI.

## 2. Methods

### 2.1 Patient Recruitment

This prospective cohort study was approved by the Research Subjects Review Board (RSRB) at the University of Rochester Medical Center (URMC). Adult patients ≥ 18 years admitted to URMC’s Cardiac Intensive Care Unit were eligible if they underwent either veno-arterial (VA, for CS or CA) or veno-venous (VV, for ARDS) ECMO therapy. Excluded were those with pre-existing neurologic conditions resulting in dependency on others for activities of daily living, acute neurologic injuries known at the time of ECMO initiation, skin injury rendering EEG impossible to obtain, facial injuries impeding DCS measurement, as well a lack of informed consent.

### 2.2 Experimental Design

A detailed description of the experimental design and instrumentation used for this study has been previously published (Dar et al., 2020, 2020). In summary, enrolled patients underwent at least two days of continuous EEG recording and two-hour concomitant daily CBF monitoring using a proprietary, lab-built DCS system. Transcranial doppler (TCD) was also obtained once daily for between 1-5 minutes; we did not display these data because TCD measurements were incomplete due to technical difficulties. All devices were approved by URMC’s Clinical Engineering Department for use in the ICU.

EEG were recorded using digital Xltek Brain Monitor EEG amplifiers (Natus, Middleton, WI, USA) with a 512 Hz sampling frequency. The gold-plated EEG electrodes were used for all patients. The Fp1 and Fp2 electrodes were placed higher than a standard position to accommodate for the DCS probe placement on the forehead. In order to accommodate for the TCD harness, the first two patients only had 11 electrodes (Fp1/2, F3/4, C3/4, P3/4, Fz, Cz, Pz). The rest of the cohort had a full montage with the exception of T7/8 electrodes to accommodate for TCD measurements; nineteen electrodes consisted of the standard parasagittal, temporal, midline and FT9/10 leads of the International 10-20 system. All electrodes were referenced to FCz. Quantitative analysis of the EEG (qEEG) was performed by Persyst™ 14 software (Persyst, Solana Beach, CA, USA). In order to perform quantitative analysis from only recording electrodes, specialized templates were constructed for patients 1 and 2 and a separate one for the rest of the cohort. The trends were sampled with artifact rejection program enabled and included FFT power for beta (>13 Hz), alpha (8-13 Hz), theta (4-7 Hz), and delta (1-3 Hz) frequency bands using a two-minute window over each measurement period for left and right hemisphere; asymmetry and relative asymmetry indices; alpha-delta ratios (ADR) for left and right anterior regions; and FFT power for Fp1-F3/Fp2-F4 channels. The EEG data were visually reviewed in accordance with ACNS standards by an epileptologist (O.S.) in their entirety on a high-resolution monitor with reconstruction of montages and adjustments of sensitivity and filtering. (Herman et al., 2015) The EEG data points comprised elements from the visual inspection in accordance with ACNS guideline (Tatum et al., 2016) and included presence of background slowing, continuity, posterior dominant rhythm (PDR), sleep elements, rhythmic and periodic patterns, epileptiform discharges, seizures, and status epilepticus. qEEG data were extracted at a 15-minute interval by E.L., O.S., and I.D. In addition, during the DCS recording, the qEEG data points were extracted more frequently (every 1-2 minutes) from the preselected EEG segments containing the least amount of interference. Recordings with a large amount of interference obscuring visual EEG analysis were not included into the analysis. The times on the devices were synchronized, and BFI data were extracted for the same two-minute time window as EEG sampling. Each band power value was converted to a percentage of the total hemispheric power for each left and right hemisphere as follows:

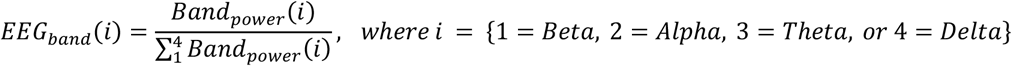

Subsequently, left and right hemispheric (Alpha+Beta)/Delta ratio (ABDR) was calculated. A Pearson correlation coefficient (R-value) was calculated between ABDR and the BFI values obtained from each single measurement time point for each hemisphere (Fig. 1). Similarly, alpha/delta ratio (ADR) was calculated for each hemisphere, but without beta band data to match with commercially available ADR concept. (Rots et al., 2016) In addition, we analyzed BFI correlation with homologous anterior quadrant ADR values. Lastly, a total spectral power for fronto-polar regions (Fp1-F3 and Fp2-F4 channels) was analyzed against BFI.

**Figure 1.**
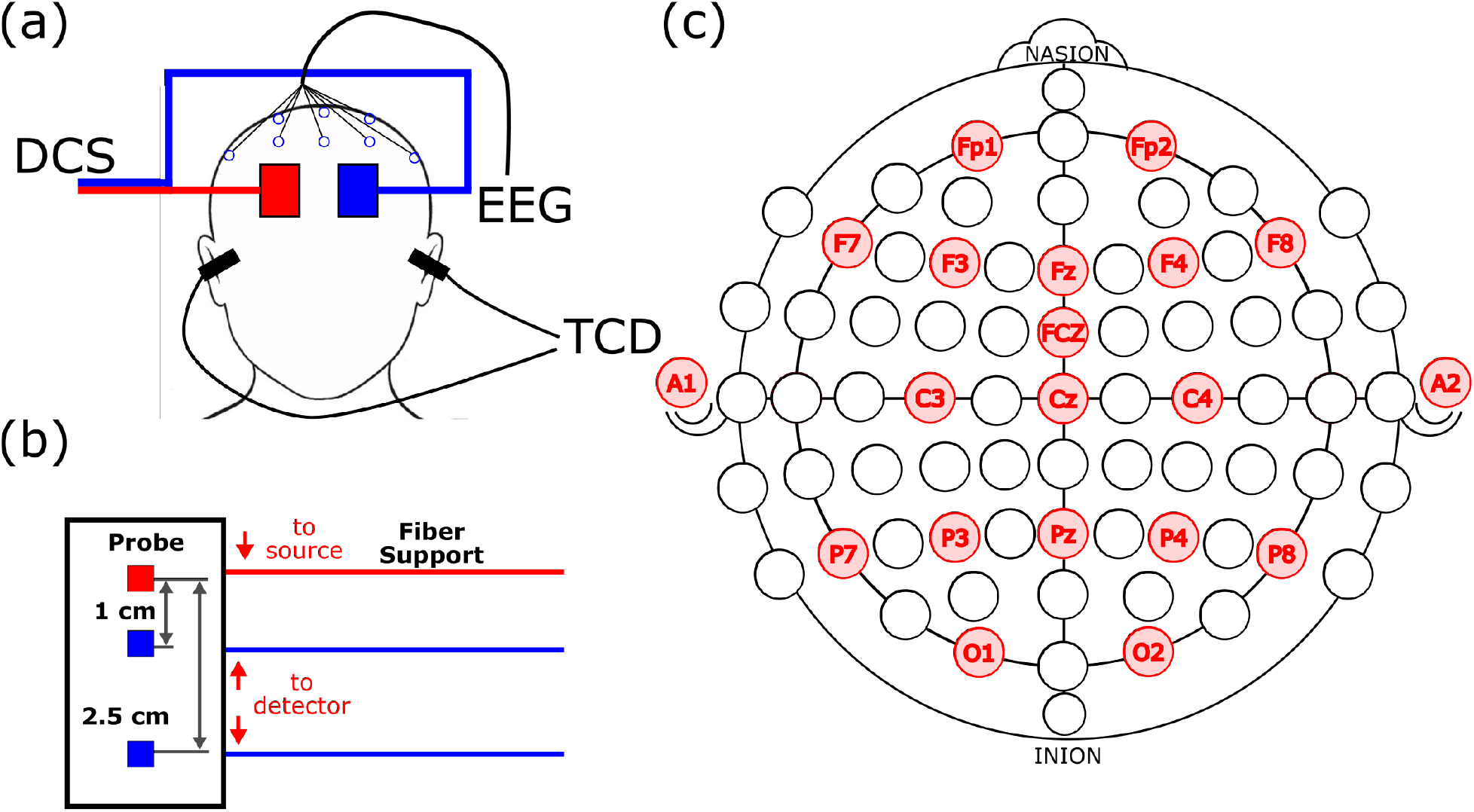
(a) The experimental setup displaying where the DCS, EEG, and TCD measurements are take. (b) Schematic of the DCS probe showing source detector separations. (c) EEG 10-20 locations where data was recorded.

The DCS system and analysis methodology used has been previously published. (Dar et al., 2020) In summary, two slim-profiled probes house fibers that connect to a 785 nm long coherence laser source and two single photon detectors at 1 cm and 2.5 cm away from the source (**figure 1**). The probes are affixed to the right and left forehead using double-sided tape and Tegaderm (both from 3M, St Paul, MN, USA). The propagated light from the source to the detector fibers are received as photon counts. Using these photon counts, an autocorrelation curve is calculated over 2 seconds at 0.25 Hz frequency alternating for each hemisphere. BFI values are then determined from these autocorrelation curves.

Neurologic examination was performed serially by the clinical team at least every four hours during sedation pauses, if clinically appropriate, and documented in the form of a total and motor subscore of Glasgow Coma Score (GCS and GCS-M respectively). (Selioutski et al., 2022) “Brain-injured” patients exhibited encephalopathy for the majority (≥50%) of days of neuromonitoring quantified as the inability to follow commands (GCS-M < 6) (Dankiewicz et al., 2021), or evidence of HIBI on CT scan before or during neuromonitoring. “Uninjured” patients were able to follow commands for the majority of neuromonitoring and did not exhibit radiographic evidence of HIBI. CT scans were obtained at the discretion of the clinical team when clinically indicated to evaluate for encephalopathy. These scans were interpreted by clinical neuroradiologists for lack of differentiation of gray versus white matter on head CT and loss of sulcal differentiation, denoting HIBI. CT scans read as normal for two patients (subjects 4 and 9) were reviewed by I.K. and T.J. for presence of HIBI quantified by the averaged gray-white ratio (GWR-AV, values <1.23 denote 100% specificity for poor outcome prediction). (Lee et al., 2016) Other demographic and clinical factors were obtained from chart review. Daily sequential organ failure assessment (SOFA) scores were calculated from daily highest severity laboratory values obtained from chart review. (Vincent et al., 1996) Outcomes data included survival to discharge and final recorded GCS score upon discharge.

### 2.3 Statistical Analysis

Patients were divided into two groups (brain-injured or uninjured). In patients without brain injury, we expect blood flow to have a positive correlation with faster frequencies and a negative correlation with slower frequencies. (Baang et al., 2022) Data was checked for normality using the Shannon-Wilks method. Using this result, demographic and clinical variables between the two groups were assessed for significant differences using the two-sample t-test if parametric or Wilcoxon rank-sum if non-parametric, or chi-square test. Statistical significance was defined as a p-value < 0.05. Statistical analysis was performed using MATLAB software.

## 3. Results

Ten patients were enrolled between 12/2019 and 7/2021, with five each in the brain-injured and uninjured groups based on the predefined criteria. Eight patients underwent VA ECMO (all for CS ± CA) while two patients with ARDS underwent VV ECMO. All patients in the uninjured group except Subject 3 were spontaneously awake and following commands for the entirety of the monitoring periods. Subject 3 was included in the uninjured group because chart review revealed ability to follow commands consistently outside neuromonitoring times and did not exhibit HIBI on CT scan. Daily GCS-motor subscores for each patient are displayed in **figure S1. Table 1** denotes demographic and clinical factors for all patients and differences between the groups. Brain-injured patients had fewer days with GCS-M=6 (24% vs. 72%, *p*=0.01) and lower GCS scores during neuromonitoring (6[6] vs. 11[8], *p*=0.01). **Table 2** lists each patient’s indication for ECMO, neuroimaging findings, and median analgosedation infusion rates during neuromonitoring. Of the two patients with CT scans read as normal per clinical neuroradiology report, Subject 4 (uninjured) was found to have a GWR-AV of 1.33 and Subject 9 (brain-injured) had a GWR-AV of 1.14.

**Table 1.** Demographic and clinical characteristics of subjects undergoing NVU monitoring. Data described as mean ± standard deviation if parametrically-distributed, median [IQR] if non-parametrically distributed. Statistically significant (p<0.05) differences between groups are denoted with an asterisk (*). ^1^ Incomplete data; two patients in encephalopathic group with unknown downtimes due to lack of documentation from transferring center. ^2^ Number of days GCS motor subscore = 6 (following commands). GCS: Glasgow coma scale. SOFA: sequential organ failure assessment. VT: ventricular tachycardia. VF: ventricular fibrillation. PEA. Pulseless electrical activity. MMM: multimodal neuromonitoring with DCS and EEG.

|  | All (n=10) | Uninjured (n=5) | Brain-injured (n=5) |
| --- | --- | --- | --- |
| Age | 45.4 ± 20.9 | 49 ± 25.3 | 41.8 ± 17.8 |
| Female (%) | 5 (50%) | 3 (60%) | 2 (40%) |
| Reason for ECMO |  |  |  |
| Cardiogenic shock (%) | 8 (80%) | 4 (80%) | 4 (80%) |
| ARDS (%) | 2 (20%) | 1 (20%) | 1 (20%) |
| Type of ECMO |  |  |  |
| VV | 2 (20%) | 1 (20%) | 1 (20%) |
| VA | 8 (80%) | 4 (80%) | 4 (80%) |
| Cardiac arrest (%) | 5 (50%) | 1 (20%) | 4 (80%) |
| Total downtime (mins)<sup>1</sup> | 15 | 30 | 7.5 |
| VT/VF (%) | 3 (30%) | 0 | 3 (60%) |
| PEA (%) | 2 (20%) | 1 (20%) | 1 (20%) |
| Length of hospitalization (days) | 42.2 ± 31.5 | 34.6 ± 30.1 | 49.8 ± 34.4 |
| Duration of ECMO (hrs) | 272.4 [167.2] | 321 [386.8] | 240.7 [120.9] |
| Duration of MMM (h:mm) | 1:59 [0:43] | 2:00 [0:47] | 1:57 [0:21] |
| Pre-ECMO SOFA | 12.5 ± 2.2 | 12.6 ± 1.9 | 12.4 ± 2.6 |
| Pre-ECMO lactate | 2.95 [8.2] | 3.2 [11.6] | 2.7 [5.1] |
| # days GCS-M=6 (%)<sup>2</sup> | 18 (46%) | 13 (72%) | 5 (24%)* |
| Median GCS | 8[8] | 11 [8] | 6 [6]* |

**Table 2.** Clinical/radiographic characteristics and analog-sedation doses of uninjured and brain-injured (shaded) groups. Analgosedation doses are displayed as average infusion rates during neuromonitoring. NM: neuromonitoring with DCS and EEG; CS: cardiogenic shock; CA: cardiac arrest; ARDS: acute respiratory distress syndrome; N/A: not available; GWR: gray-white ratio, values <1.23 denote 100% specificity for poor outcome prediction. ^1^“Awake” is defined as able to follow commands. ^2^In mcg/kg/hr. ^3^In mg/hr. ^4^In mcg/hr. ^5^Neurologic exam denoted for monitoring days during which patient was not chemically paralyzed.

| Patient | Indication | # NM days | % NM days awake<sup>1</sup> | Imaging (GWR) | Sedation |  |  | Analgesia |  |
| --- | --- | --- | --- | --- | --- | --- | --- | --- | --- |
|  |  |  |  |  | Propofol<sup>2</sup> | Midaz-olam<sup>3</sup> | Dexmed-etomidine<sup>2</sup> | Fentanyl<sup>4</sup> | Hydro-morphone<sup>3</sup> |
| 1 | CS + CA | 3 | 100 % | N/A | 0 | 0 | 0 | 58.7 | 0 |
| 2 | CS | 4 | 100 % | N/A | 0 | 0 | 0 | 25 | 0 |
| 3 | CS | 2 | 25 % | N/A | 15 | 0 | 0.8 | 0 | 1.8 |
| 4 | CS | 3 | 100 % | Normal (GWR 1.33) | 0 | 0 | 0 | 0 | 0 |
| 10 | ARDS | 6 | 100%<sup>5</sup> | N/A | 35 | 8.7 | 1.0 | 0 | 6 |
| 5 | CS + CA | 7 | 29 % | HIBI | 0 | 0 | 0.4 | 100 | 0 |
| 7 | ARDS | 5 | 20 %<sup>5</sup> | HIBI | 0 | 2 | 0 | 187.6 | 0 |
| 9 | CS + CA | 2 | 0 % | Normal (GWR 1.14) | 0 | .5 | 0 | 37.5 | 0 |
| 11 | CS + CA | 2 | 0 % | HIBI | 17.6 | 0 | 0 | 90.4 | 0 |
| 12 | CS + CA | 5 | 29 % | N/A | 0 | 0 | 0 | 0 | 0 |

**Table 3** displays EEG features for uninjured and brain-injured patients. No brain-injured patients exhibited anterior-posterior gradients, sleep architecture, or PDR, while the majority of uninjured patients had these features. All patients exhibited continuous, slow backgrounds for the majority of their recordings and none had epileptiform discharges, rhythmic or periodic patterns, seizures, or status epilepticus.

**Table 3.** cEEG features for uninjured and brain-injured (shaded) groups. (+): present for the majority (>50%) of the recording, (-): absent for majority of recording. ^1^In hours:minutes, excluding uninterpretable portions. ^2^Average alpha delta ratio (ADR) listed as [right hemisphere; left hemisphere]. ^3^Defined as maximum – minimum ADR for each patient [right hemisphere; left hemisphere]. GDS: generalized diffuse slowing; FIRDA: frontal intermittent rhythmic delta activity.

| Patient | cEEG<br>durat-<br>ion<sup>1</sup> | Average<br>ADR<sup>2</sup><br>[R/L] | ADR range<br>[R/L]<sup>3</sup> | Back-<br>ground | AP Gradient | Reactivity | Variability | Sleep | PDR | Total<br>features<br>present |
| --- | --- | --- | --- | --- | --- | --- | --- | --- | --- | --- |
| 1 | 8:31 | [0.88; 0.86] | [1.11; 1.14] | FIRDA | + | + | + | + | + | 5 |
| 2 | 9:15 | [0.48; 0.49] | [0.40; 0.37] | GDS | + | + | + | + | + | 5 |
| 3 | 5:56 | [0.20; 0.21] | [0.17; 0.17] | GDS | - | + | - | - | - | 1 |
| 4 | 5:02 | [0.26; 0.27] | [0.48; 0.56] | GDS | + | + | - | + | + | 4 |
| 10 | 13:27 | [0.41; 0.40] | [0.44; 0.46] | GDS | - | + | + | + | - | 3 |
| *Mean* | *8:26* | *[0.45; 0.45]* | *[0.52; 0.54]* | *Total (%)* | *60* | *100* | *60* | *80* | *60* |  |
| 5 | 20:31 | [0.38; 0.40] | [0.32; 0.34] | GDS | - | + | - | - | - | 1 |
| 7 | 9:36 | [0.67; 0.57] | [1.77; 1.61] | GDS | - | - | + | - | - | 1 |
| 9 | 4:06 | [0.17; 0.17] | [0.13; 0.20] | GDS | - | - | - | - | - | 0 |
| 11 | 3:51 | [0.48; 0.52] | [1.60; 1.45] | GDS | - | - | - | - | - | 0 |
| 12 | 9:24 | [0.26; 0.28] | [0.31; 0.33] | GDS | - | + | + | - | - | 2 |
| *Mean* | *9:30* | *[0.39; 0.39]* | *[0.83; 0.79]* | *Total (%)* | *0* | *40* | *40* | *0* | *0* |  |

ABDR, ADR, and BFI data from a total of 39 monitoring sessions were analyzed. The median duration of each DCS monitoring session was 1 hour 59 minutes (IQR 49 minutes) with an average of 3.9 sessions per patient. No differences in monitoring duration or number of sessions were noted between groups. Correlation for each EEG power band versus BFI is displayed in **table S1**, and mean power for each EEG frequency band for both groups is listed in **table S2. Figure 2** displays correlation plots of ABDR versus BFI for each patient with data for all neuromonitoring days included. Brain-injured patients were found to have significantly lower correlation between ABDR and BFI than uninjured patients (left: 0.52 ± 0.38 vs. 0.05 ± 0.14, *p*=0.03, right: 0.39 ± 0.42 vs. -0.12 ± 0.24, *p*=0.04, **figure 3a**). Correlations between hemispheric ADR and BFI were also calculated and plotted, also revealing lower correlations in brain-injured patients (**figures S2 and 3b**, left: 0.51 ± 0.37 vs. 0.14 ± 0.13, *p*=0.07, right: 0.47 ± 0.43 vs. -0.06 ± 0.22, *p*=0.04). **Figure S3** displays anterior quadrant-specific ADR and BFI correlation for each patient with similar differences between brain-injured and uninjured patients (left: 0.52 ± 0.39 vs. 0.11 ± 0.20, *p*=0.07, right: 0.50 ± 0.47 vs. -0.13 ± 0.24, *p*=0.03). **Figure S4** displays correlation between fronto-polar spectral power and BFI; no significant differences were noted between groups.

**Figure 2.**
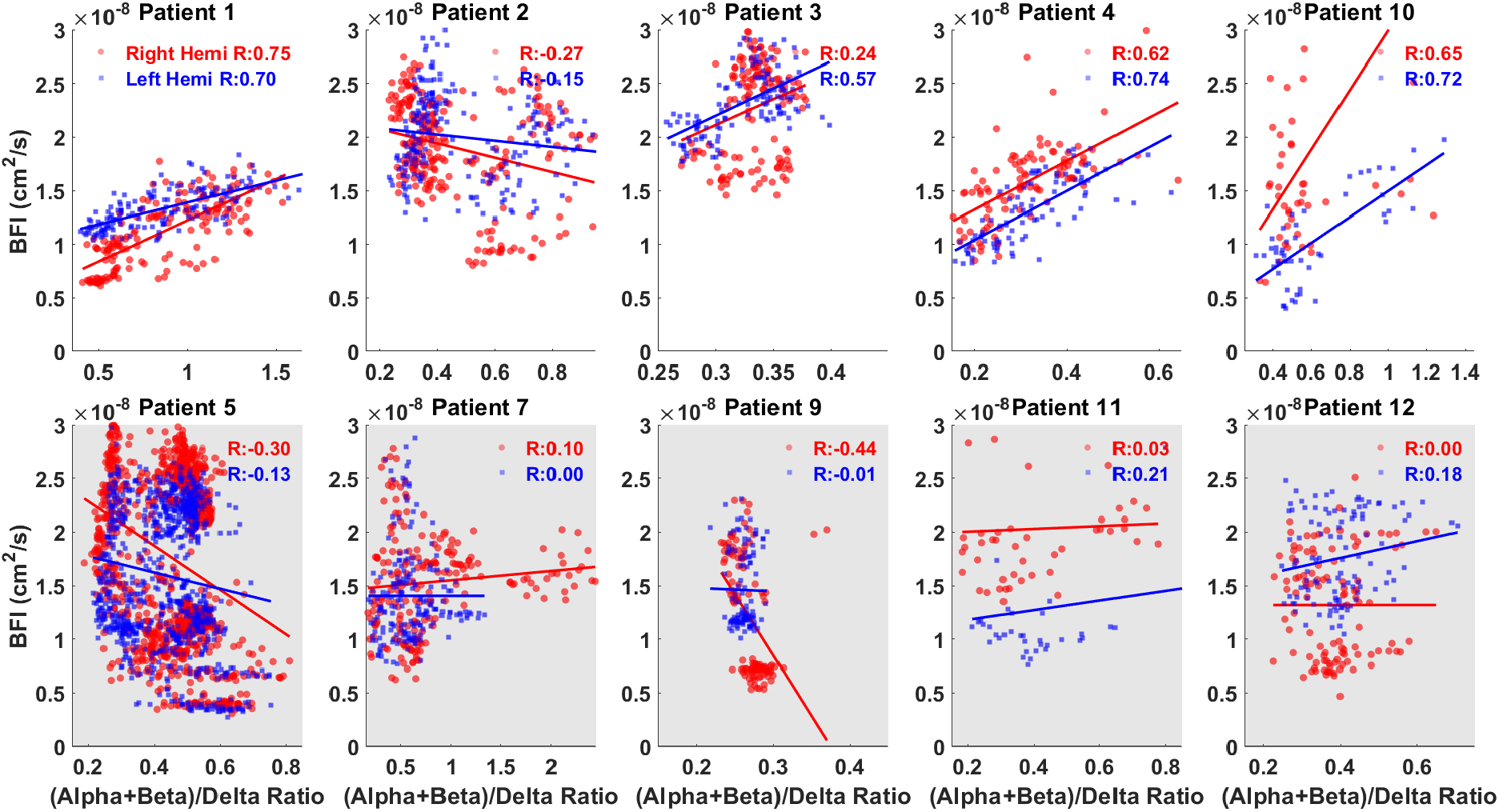
ABDR vs. BFI plots for all patients. Unshaded plots indicate uninjured patients, while shaded plots indicate brain-injured patients. Red dots indicate right hemisphere data, blue dots indicate left hemisphere data.

**Figure 3.**
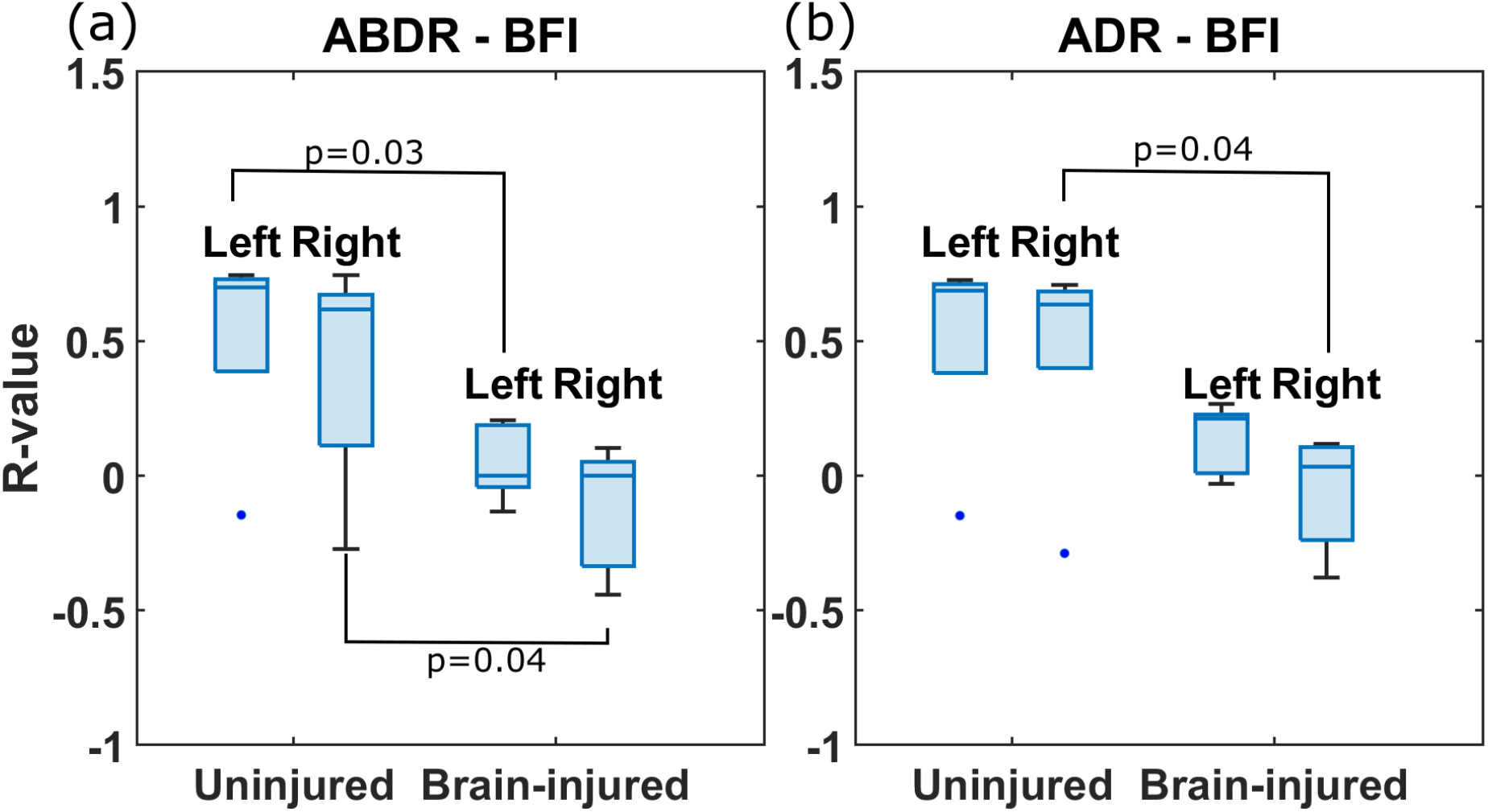
Box plots comparing hemispheric (a) ABDR vs. BFI and (b) ADR vs. BFI for uninjured and brain-injured groups.

Each patient’s outcome is displayed in **table 4**. Three patients survived in either group, and a higher number of patients were awake and following commands by time of discharge in the uninjured group than the brain-injured group (80% vs. 40%, *p*=0.20).

**Table 4.** Outcomes for uninjured and brain-injured (shaded) groups. ^1^Last recorded Glasgow Coma Score (GCS) and GCS motor subscore (GCS-M) prior to discharge or decision to withdraw care. ^2^Patient died of hemorrhagic shock from tracheostomy, in addition to refractory ARDS and ECMO cannula-induced lower leg ischemia. ARF: acute rehabilitation facility

| Patient | Survived ECMO | Survived to discharge | Care withdrawn | Cause of death | Last GCS score<sup>1</sup> | Last GCS-M<sup>1</sup> | Where was patient discharged to? |
| --- | --- | --- | --- | --- | --- | --- | --- |
| 1 | Yes | Yes |  |  | 15 | 6 | Nursing home/SNF |
| 2 | No | No | Yes | Sepsis | 11 | 6 |  |
| 3 | No | No | Yes | Hemorrhage<sup>2</sup> | 3 | 1 |  |
| 4 | Yes | Yes |  |  | 15 | 6 | Home without therapy |
| 10 | Yes | Yes |  |  | 15 | 6 | ARF |
| 5 | Yes | Yes |  |  | 15 | 6 | Nursing home/SNF |
| 7 | Yes | Yes |  |  | 15 | 6 | ARF |
| 9 | No | No | Yes | Sepsis | 3 | 1 |  |
| 11 | Yes | Yes |  |  | 6 | 1 | ARF |
| 12 | Yes | No | Yes | Cardiovascular | 8 | 4 |  |

## 4. Discussion

This study paired two noninvasive modalities, quantitative EEG assessing cortical electrical activity and DCS assessing cortical CBF, to evaluate the correlation between neuronal function and perfusion in adults undergoing ECMO. Brain-injured patients exhibited significantly lower correlation between BFI and ABDR or ADR than uninjured patients. One possible explanation for this decreased correlation is due to NVU dysfunction. Our study population experienced severe ARDS or cardiogenic shock with or without cardiac arrest, predisposing them to HIBI. Numerous studies of patients with HIBI link elevated blood biomarkers of damaged neurons (neuron-specific enolase, NSE), axons (neurofilament light, NFL), and astrocytes (glial fibrillary acid protein, GFAP) to poor neurologic outcomes. (Diaz-Arrastia et al., 2020, Hoiland et al., 2022, Stammet, 2017) The NVU is comprised of these cell types together with pericytes and capillary endothelium, implicating it as the site of injury in patients with HIBI. (Iadecola, 2017) Under normal circumstances, neuronal activity contributes to the endothelial production of vasodilators including nitric oxide, prostanoids, and adenosine, all of which act to regulate cerebral blood flow. (Filosa et al., 2006, Iadecola et al., 1993, Lecrux and Hamel, 2016) Whole brain ischemia leads to an oxygen diffusion limitation even after perfusion has been restored, contributing to ongoing ischemia termed the “no-reflow” phenomenon. (Li et al., 2019, Sekhon et al., 2020) This ongoing ischemia causes a metabolic crisis for neurons, decreasing their ability to stimulate vasoactive substances ultimately contributing to the loss of cerebrovascular autoregulation. (Bundo et al., 2002, Dietrich et al., 1986, Girouard and Iadecola, 2006) In the absence of other causes of brain injury, we assume HIBI is the underlying cause of brain injury in our population, and these processes may explain why they experienced decreased ABDR-BFI correlation.

While NVU structure and function have garnered significant attention in recent studies, descriptions of *in vivo* NVU monitoring in humans are in their early stages. Studies have previously used neuroimaging modalities such as fMRI and CT perfusion combined with EEG to assess NVU function. (Ajcevic et al., 2021, Jafarian et al., 2020) Though imaging-based NVU assessment provides spatial resolution, it does not allow for continuous monitoring of dynamic processes which are often seen in acutely brain-injured populations. Invasive intracranial monitoring modalities offer data on cerebral metabolism, perfusion, and tissue oxygenation, but this can be dangerous in ECMO patients due to the use of anticoagulation and ECMO-induced clotting factor disruption. (Millar et al., 2016, Thomas et al., 2018) By using DCS, our methodology takes advantage of optical technology to measure CBF noninvasively and continuously at the bedside and can easily be performed in conjunction with EEG.

Previous studies have demonstrated the utility of qEEG to calculate a ratio of fast versus slow EEG power bands to detect cerebral ischemia due to cardiac arrest (Wiley et al., 2018) and subarachnoid hemorrhage (Yu et al., 2019). The distribution of EEG frequencies depends on the patient’s sleep-wake state, functional neuronal status, adequacy of blood supply, and medication-induced sedation. A calm, awake, neurologically-intact patient with eyes closed has a PDR, noted by alpha-range activity over the occipital regions and marked beta activity at 14-16 Hz in the fronto-central regions. (Ebersole and Pedley, 2003) Delta frequencies dominate the EEG background during deep sleep, analgosedation use, or brain injury. (Tatum IV, 2021) EEG changes occur in tandem with alterations in CBF. These changes are either physiologic, such as CBF reduction during deep sleep due to decreased cerebral synaptic activity (Madsen and Vorstrup, 1991), or pathological, as in case of transient ischemic attack or stroke when reduced blood supply leads to loss of faster frequencies and increased slowing. (Foreman and Claassen, 2012, Gottlibe et al., 2020)

As opposed to previous studies which only include alpha frequency activity, we chose to combine alpha and beta frequencies in our analysis for a number of reasons specific to our population and monitoring paradigm. First, beta frequencies are predominantly noted anteriorly in the awake, uninjured brain as mentioned above. We measured CBF in the frontal lobes using DCS. Thus, including widely distributed beta frequencies allows a more accurate measurement of all EEG frequencies in regions relevant to monitored CBF. Second, beta activity is increased by benzodiazepines (Fisch, 1999), which are commonly used in critically-ill patients to maintain ventilator compliance and prevent accidental cannula dislodgement. (Barr et al., 2013, Burcham et al., 2017) ECMO can prolong the effect of these drugs by increasing their volume of distribution and sequestering them throughout the circuit. (Burcham et al., 2017) The presence of cerebral beta activity is highly dependent on normal cortical function and its presence is a good prognostic sign, whereas depression of the beta activity is a reliable indicator of abnormality and should not be disregarded. (Ebersole and Pedley, 2003, Fisch, 1999) Notably, ABDR and ADR both had higher correlations with CBF in uninjured patients than in brain-injured ones, with ABDR-BFI achieving statistical significance for both hemispheres. One possible reason we did not see a significant difference in correlation patterns between ABDR-BFI (**figure 2)** and ADR-BFI (**figure S2**) is that benzodiazepines were rarely used in our cohort (**table 2**). Notably, the brain-injured group did receive fentanyl infusion more often and at higher doses than the uninjured group. Fentanyl can decrease EEG frequency but not cerebral perfusion, and could have contributed to decreased ABDR-BFI correlation. However, our findings were consistent in uninjured patients who received fentanyl (subjects 1 and 2) and a brain-injured patient who did not (subject 12). Our groups were not adequately powered to determine whether ABDR-BFI correlations associated with differences in mortality, withdrawal of care, or final recorded GCS scores.

Anterior region-specific ADR-BFI correlation analysis (**figure S3**) appeared grossly similar to hemispheric ADR-BFI analysis with marginal lower correlation in brain-injured patients. This lack of regional difference may support our use of whole-hemisphere EEG power data to examine an extrapolated correlation even though CBF is only being monitored at the frontal region. It also supports the addition of beta to alpha band power to improve frontal electrographic sensitivity in our correlation analysis. The lack of correlation between fronto-polar EEG and BFI was unexpected as we anticipated there to be a correlation in uninjured patients, who should have higher spectral beta power in the frontal regions. However, our sample was not powered to detect this difference, and limiting EEG analysis to two lead pairs (Fp1-F3 and Fp2-F4) may be confounded by overlap in electrical fields with one another. Finally, we moved Fp1 and Fp2 leads superiorly by one centimeter closer to F3/F4, which could hamper spectral power calculations.

There are several limitations to our study. First, we acknowledge there may be alternate explanations for the lack of correlation between ABDR and BFI in brain-injured patients. Rather than at the level of the NVU and capillary bed, the correlation between ABDR and BFI could be driven by cerebrovascular autoregulation at the arterial level. This mechanism maintains cerebral perfusion at a steady state in response to changes in systemic blood pressure. (van den Brule et al., 2018) Cerebrovascular autoregulation is noted to be altered after cardiac arrest (Laurikkala et al., 2021, Sekhon et al., 2019) and in those with non-pulsatile blood flow (Veraar et al., 2019), rendering cerebral perfusion more sensitive to systemic hemodynamics. This is especially relevant in patients undergoing VA ECMO, which is inherently non-pulsatile but pulsatility varies with cardiac contractility. Furthermore, it is unclear whether microvascular perfusion regulation via the NVU or macrovascular regulation via systemic hemodynamics work independently or in conjunction. We did not measure the effect of cerebral autoregulation or pulsatility in this study because of the heterogeneity of our patient population; not all patients underwent VA ECMO or had pulmonary artery catheters to measure high-fidelity hemodynamics. The methodology was based on a 2-minute window of EEG averaging. Lastly, the ABDR-BFI correlation only measures perfusion-associated electrographic activity without delineating the cause of pathophysiology.

The heterogeneity of our study population is a second limitation to our study design. In trying to capture as many patients with the potential for HIBI in as short a time as possible, we enrolled patients who had experienced ARDS, cardiogenic shock, or cardiac arrest. All three of these pathophysiologies can cause various types of brain injury to varying degrees. (Huang et al., 2021, Khan et al., 2021) In addition to this, VA and VV ECMO can cause varying, uncertain degrees of brain injury. (Millar et al., 2016) This limits the causal relationship between our findings and their underlying pathophysiology. However, our study’s aim was to describe the relationship between BFI and EEG frequencies as a marker of neuronal dysfunction in patients with HIBI regardless of underlying cause. Third, the classification of “brain-injured” and “uninjured” was dependent on clinical exam (GCS) during ECMO, which is often obscured by analgosedation. We used this classification criteria because it is our institution’s standard practice for neurologic evaluation and drives clinical decision-making for subsequent neurologic workup. Fourth, we acknowledge the small sample size used in our study, limiting the generalizability of our findings. Future studies will include larger, more homogenous populations to establish inter-patient validity. Finally, we acknowledge that although the first two patients (one in each group) had fewer EEG electrodes compared to the rest of the group, they were included into the analysis. This decision was based on the observation of all expected EEG background findings in patient #1. All future patients will have the maximal EEG electrode set.

The TCD probes and its head frame necessitated a limited EEG montage for some patients as described in section 2.2. Given that patients with limited EEG montages were evenly distributed among two groups we chose not to exclude them from the analysis. However, limited EEG input could impact hemispheric ABDR values. In the future, full EEG arrays will be used.

Monitoring correlations between CBF and neuronal function noninvasively can have important downstream applications for therapeutics research and clinical practice. Future studies can establish a link between NVU dysfunction and ABDR-BFI correlation by measuring NVU damage using blood biomarkers (e.g., NFL, NSE, GFAP). If confirmed as a marker of NVU dysfunction, our paradigm can inform efficacy of future therapies directed at neuroglial or endothelial injury in patients with HIBI. The noninvasive nature of both DCS and EEG opens a number of populations, such as coagulopathic ECMO patients, to investigations of multimodal neuromonitoring. Finally, the ABDR-BFI correlation may also provide a valuable marker of neuroprognostication in comatose patients, or in those who are otherwise difficult to examine due to analgosedation.

## 5. Conclusion

Brain-injured ECMO patients with severe ARDS, cardiogenic shock, or refractory cardiac arrest exhibited significantly less correlation between CBF and ABDR than uninjured patients. The combined use of DCS and quantitative EEG holds promise as a noninvasive, continuous, multimodal indicator of brain injury.

## Data Availability

All data produced in the present study are available upon reasonable request to the authors

## Funding

This work was supported by the University of Rochester 2019 University Research Award.

## In Online Supplement

**Figure S1**. Glasgow Coma Score Motor (GCS-M) subscores for each patient during neuromonitoring (red) and highest GCS-M score (blue) for each day of neuromonitoring.

**Figure S2**. Hemispheric ADR vs. BFI plots for each patient. Unshaded plots indicate uninjured patients, while shaded plots indicate brain-injured patients. Red dots indicate right hemisphere data, blue dots indicate left hemisphere data.

**Figure S3**. Anterior quadrant ADR vs. BFI plots for each patient. Unshaded plots indicate uninjured patients, while shaded plots indicate brain-injured patients. Red dots indicate right hemisphere data, blue dots indicate left hemisphere data.

**Figure S4**. Box plots comparing hemispheric anterior ADR vs. BFI for uninjured and brain-injured groups

**Figure S5**. Fronto-polar total spectral power vs. BFI plotted for each patient. Unshaded plots indicate uninjured patients, while shaded plots indicate brain-injured patients. Red dots indicate right hemisphere data, blue dots indicate left hemisphere data.

## Notes

### Competing Interest Statement

The authors have declared no competing interest.

### Funding Statement

This study was supported by the University of Rochester 2019 University Research Award.

### Author Declarations

The Research Subjects Review Board of the University of Rochester Medical Center gave ethical approval for this work

