## Supplementary material for "Correlations Between EEG Spectral Power and Cerebral Perfusion in Adults Undergoing Extracorporeal Membrane Oxygenation": Online Supplement

**Online Supplemental Material**

| | Right Hemisphere R-value | | | | Left Hemisphere R-value | | | |
|---|---|---|---|---|---|---|---|---|
| Patient | Alpha | Beta | Delta | Theta | Alpha | Beta | Delta | Theta |
| 1 | 0.74 | 0.80 | -0.75 | -0.55 | 0.82 | 0.80 | -0.72 | -0.66 |
| 2 | -0.31 | -0.22 | 0.34 | -0.38 | -0.11 | -0.08 | 0.15 | -0.22 |
| 3 | 0.74 | -0.55 | -0.22 | 0.04 | 0.56 | 0.56 | -0.53 | 0.13 |
| 4 | 0.20 | 0.53 | -0.22 | 0.03 | 0.11 | 0.54 | -0.25 | 0.19 |
| 10 | 0.62 | 0.62 | -0.62 | 0.46 | 0.62 | 0.63 | -0.70 | 0.65 |
| 5 | -0.23 | -0.56 | 0.09 | 0.18 | -0.14 | -0.47 | -0.02 | 0.29 |
| 7 | 0.15 | 0.01 | -0.09 | -0.07 | -0.02 | -0.22 | 0.12 | -0.10 |
| 9 | -0.46 | -0.58 | 0.22 | 0.27 | 0.07 | -0.53 | -0.38 | 0.52 |
| 11 | 0.03 | 0.03 | -0.02 | 0.00 | 0.18 | 0.14 | -0.21 | 0.29 |
| 12 | 0.17 | -0.43 | 0.11 | -0.35 | 0.31 | -0.21 | -0.09 | -0.18 |

**Table S1.** Correlation (R-values) for all EEG power bands versus BFI. Brain-injured patients are shaded.

| | Uninjured | Brain-injured | *p* | Uninjured (range) | Brain-injured (range) | *p* |
|---|---|---|---|---|---|---|
| *Right Hemisphere* | | | | | | |
| Alpha | 1.19 ± 0.21 | 1.21 ± 1.33 | 0.84 | 1.09 ± 0.29 | 1.88 ± 3.70 | 0.31 |
| Beta | 0.67 ± 0.09 | 0.69 ± 0.84 | 0.84 | 0.55 ± 0.18 | 1.20 ± 1.78 | 0.15 |
| Theta | 4.39 ± 2.01 | 3.91 ± 3.36 | 0.55 | 4.43 ± 1.86 | 8.04 ± 4.36 | 0.42 |
| Delta | 1.83 ± 0.70 | 1.76 ± 1.20 | 0.31 | 1.79 ± 0.78 | 2.88 ± 2.49 | 0.69 |
| *Left Hemisphere* | | | | | | |
| Alpha | 1.26 ± 0.44 | 1.34 ± 1.16 | 1.00 | 1.40 ± 1.10 | 2.38 ± 4.28 | 0.42 |
| Beta | 0.67 ± 0.23 | 0.78 ± 0.67 | 0.84 | 0.66 ± 1.14 | 1.67 ± 2.05 | 0.42 |
| Theta | 4.76 ± 1.79 | 3.57 ± 3.57 | 0.55 | 6.33 ± 4.64 | 7.42 ± 5.94 | 0.84 |
| Delta | 1.95 ± 0.85 | 1.54 ± 1.41 | 0.55 | 2.43 ± 1.96 | 2.75 ± 2.58 | 0.84 |

**Table S2.** Hemispheric median power and range (in microvolts$^2$/Hz) for each EEG frequency band in uninjured and brain-injured groups.

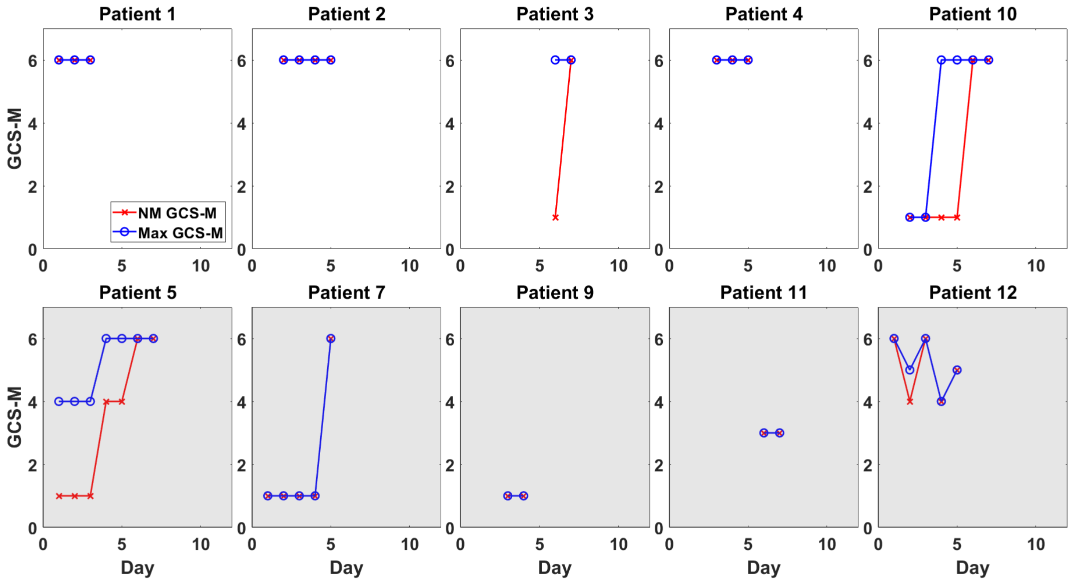


**Figure S1.** Glasgow Coma Score Motor (GCS-M) subscores for each patient during neuromonitoring (red) and highest GCS-M score (blue) for each day of neuromonitoring.

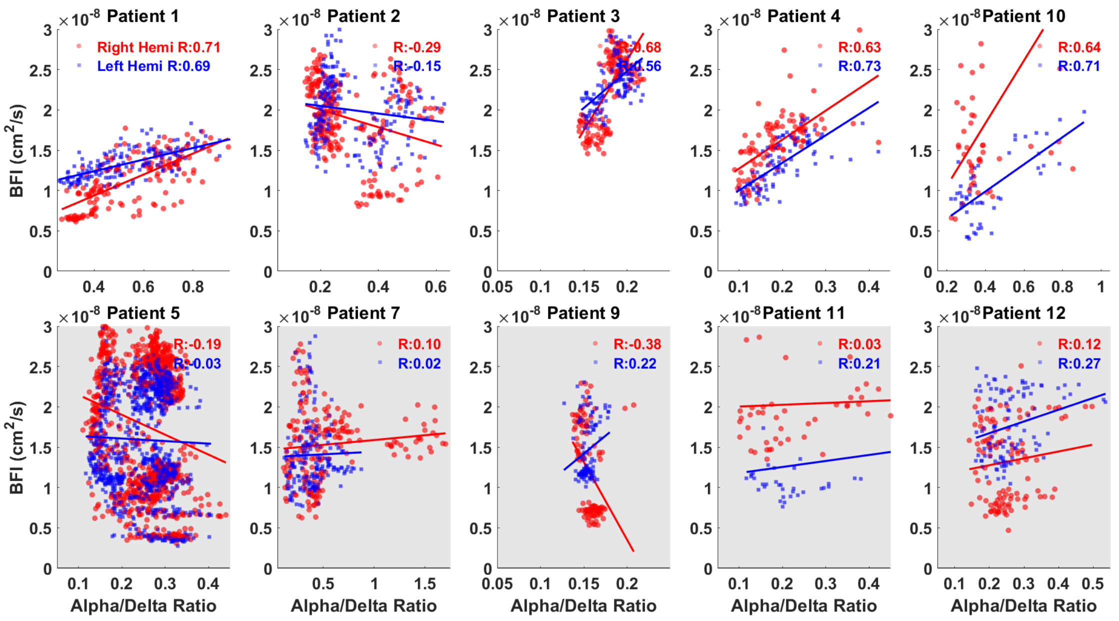


**Figure S2.** Hemispheric ADR vs. BFI plots for each patient. Unshaded plots indicate uninjured patients, while shaded plots indicate brain-injured patients. Red dots indicate right hemisphere data, blue dots indicate left hemisphere data.

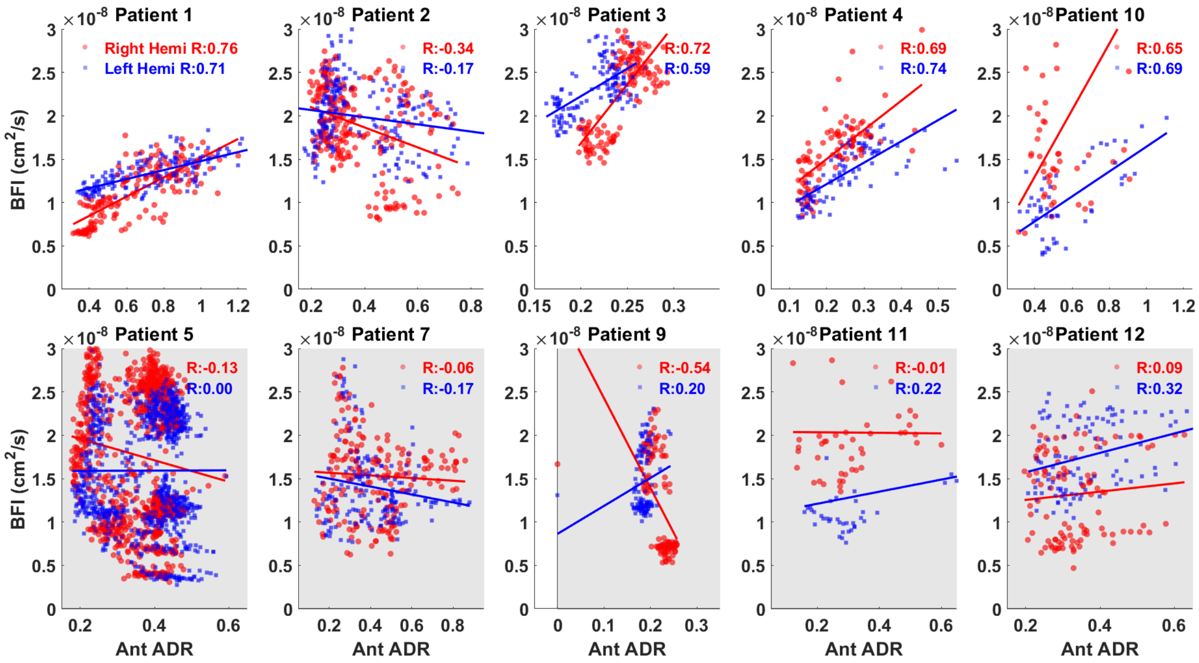


**Figure S3.** Anterior quadrant ADR vs. BFI plots for each patient. Unshaded plots indicate uninjured patients, while shaded plots indicate brain-injured patients. Red dots indicate right hemisphere data, blue dots indicate left hemisphere data.

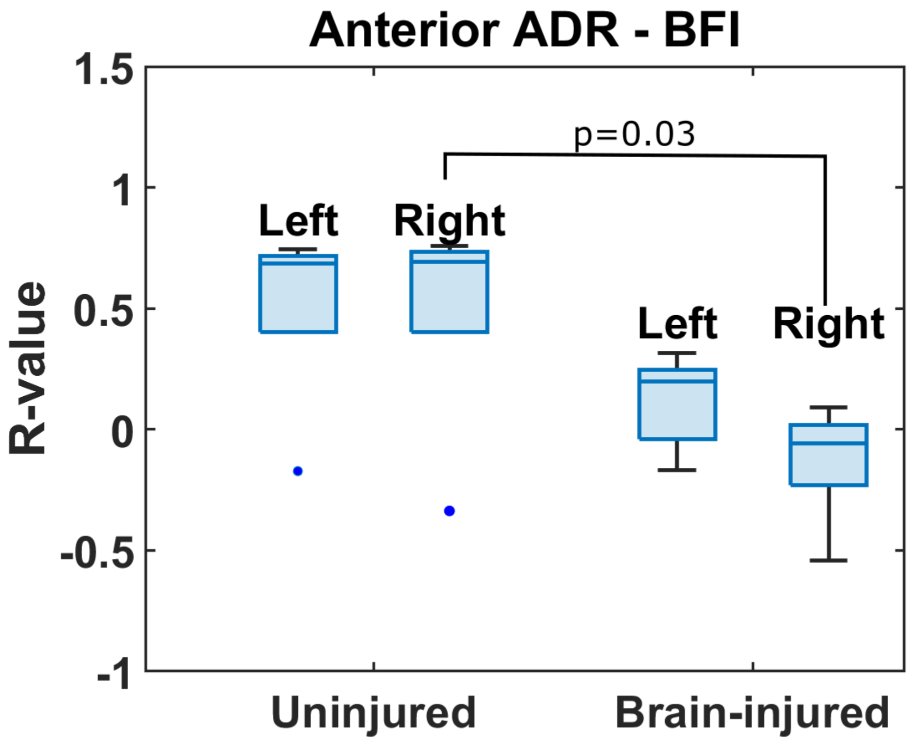


**Figure S4.** Box plots comparing hemispheric anterior ADR vs. BFI correlation for uninjured and brain-injured groups

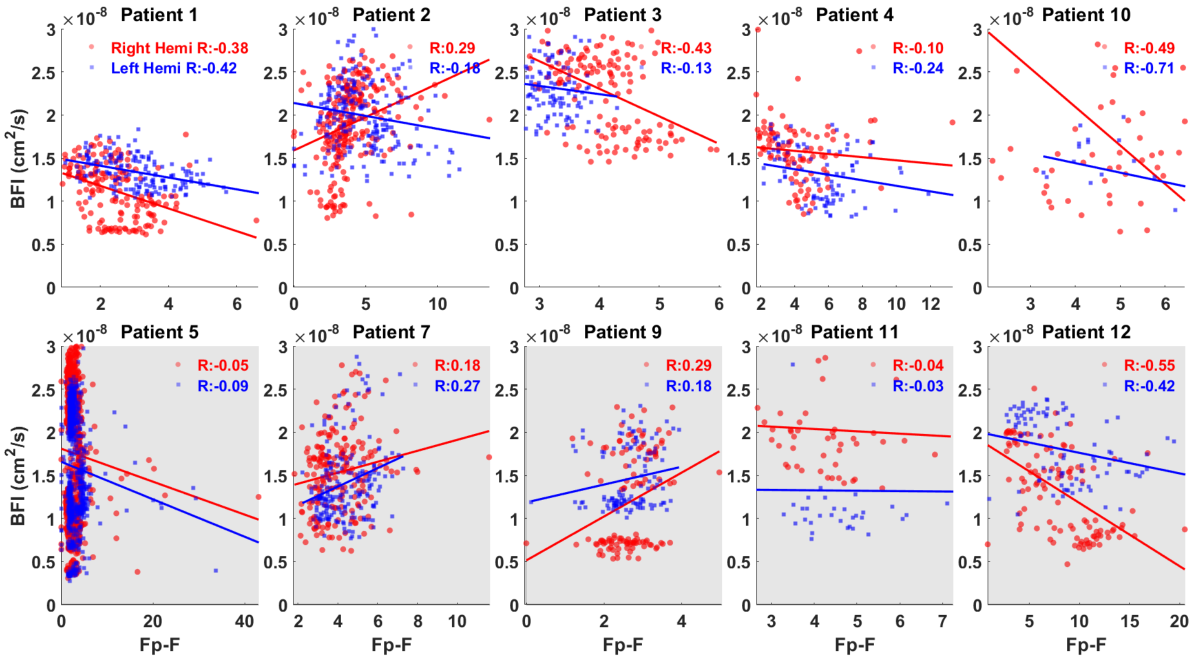


**Figure S5.** Fronto-polar total spectral power vs. BFI plotted for each patient. Unshaded plots indicate uninjured patients, while shaded plots indicate brain-injured patients. Red dots indicate right hemisphere data, blue dots indicate left hemisphere data.
